# Association of national Bacille Calmette-Guérin vaccination policy with COVID-19 epidemiology: an ecological study in 78 countries

**DOI:** 10.1101/2020.05.13.20100156

**Authors:** Norifumi Kuratani

## Abstract

A possible association between national Bacille Calmette-Guérin (BCG) vaccination policy and lower COVID-19 incidence has been suggested in some preprint papers. Using publicly accessible databases, I explored associations of national BCG vaccination policy with COVID-19 epidemiology in 78 countries. Data collection was conducted from April 25 to May 5, 2020. I compared countries that have a current universal BCG vaccination policy (BCG countries), with countries that currently lack such a policy (non-BCG countries). The mixed effect model revealed national BCG policy decreases in the country-specific risk of death by COVID-19, correspond to odds ratio of 0.446 (95% confidence interval 0.323 - 0.614, P=1×10^−5^). In BCG countries, the case increase rate was attenuated marginally by 25.4% (95% confidence interval 3.0 to 42.7, P=0.029) as compared with those of the non-BCG countries. Although the protective mechanism of BCG vaccination against COVID-19 remains unknown, further laboratory and clinical research should be warranted.

## Introduction

A possible association between national Bacille Calmette-Guérin (BCG) vaccination policy and lower COVID-19 incidence has been suggested in some preprint papers.^1,2,3^ Given the seriousness of the worldwide COVID-19 pandemic and the lack of an effective mitigation strategy, the protective effect of BCG on COVID-19 epidemic is a matter of great interest. Although an ecological study can be confounded by various biases and an ecological fallacy is always concern to understand the results, the findings can provide the evidence to conduct further prospective clinical researches. I examined the association between national BCG vaccination policy and the case fatality rate of COVID-19 in 78 countries. I also explored the relationship between BCG vaccination policy and the speed of COVID-19 increase within the countries.

## Methods

Data for the analysis were extracted from publicly accessible databases on websites.^4^ Data collection was conducted from April 25 to May 5, 2020. Inclusion criteria of the countries analyzed were: 1) defined by the World Bank as an upper-income or upper-middle-income country,^5^ and 2) country for which the necessary data were available. Countries with less than 100 confirmed cases were excluded from the analysis. I compared countries that have a current universal BCG vaccination policy (BCG countries), with countries that currently lack such a policy (non-BCG countries).^6^

I focused two outcome measures to evaluate the impact of national BCG policy on COVID-19 epidemiology. In this study, the case fatality rate (CFR) was defined as the proportion of reported number of deaths due to COVID-19 compared to the total confirmed cases on April 25, 2020. The CFRs of the countries were stratified by BCG policy and were combined by fixed or random effects models to estimate the pooled CFR with 95% confidence intervals (95% CI). A mixed effect logistic regression model was constructed to calculate a country-specific odds ratio for the effect of national BCG policy on the CFR.

I also tried to determine whether the speed of COVID-19 case increase within the countries was associated with their national BCG policy. I summarized the number of weekly new diagnosed COVID-19 patients in the initial 8 weeks of the epidemic. Because the beginning of the COVID-19 epidemic varied by country, I defined the Day 1 as the day the first confirmed COVID-19 patients was reported in the respective country. Using a generalized estimating equation (GEE) with a log-linear link function, I estimated if the slopes of case numbers were different between BCG and non-BCG countries. In the GEE analysis, the population of the country was incorporated as an offset term for adjustment, and robust “sandwich” variance estimators were used.

I did all analyses in R (version 3.6.3, “metaprop” for the CFR pooling, “lme4” for mixed effect model, “geepack” for GEE). I considered *P* values of less than 0.05 to be significant.

## Results

A total of 78 countries (BCG countries: n=54; non-BCG countries: n= 24) met for the inclusion criteria for this study. The total number of deaths in the 78 countries was 198,200 (on April 25, 2020) and this accounted for 97.5% of world COVID-19 mortalities to that date.

As Figure 1 illustrates, there were significant heterogeneities in the CFR among the countries analyzed. The pooled CFR stratified by national BCG policy was significantly lower mortality in the BCG countries than in the non-BCG countries (Figure 1). The mixed effect model revealed national BCG policy decreases in the country-specific risk of death correspond to odds ratio of 0.446 (95% CI 0.323 - 0.614, P=1×10^−5^).

**Figure 1:**
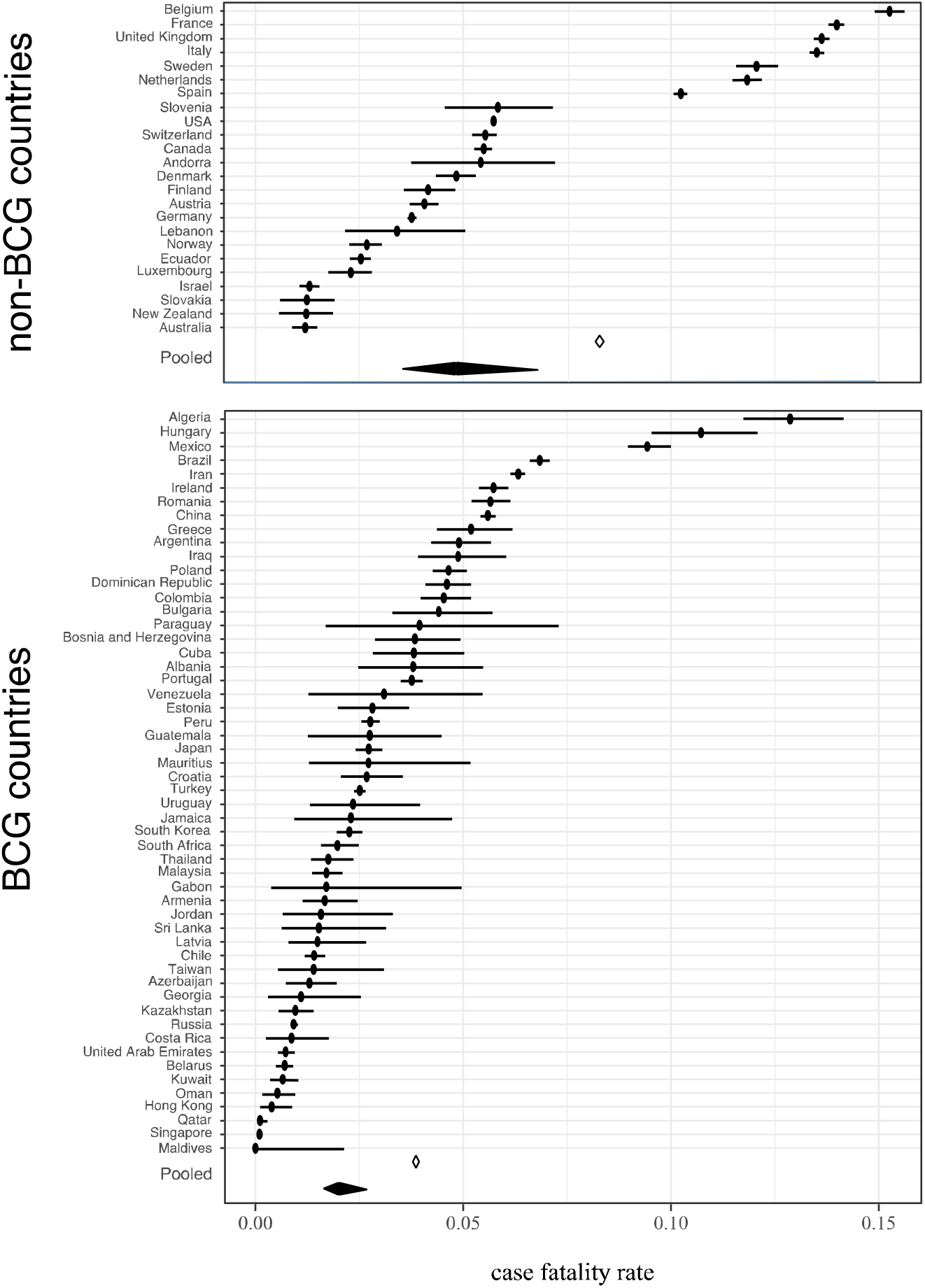
Forest plot of the COVID-19 case fatality rate in 78 countries. The black circles indicate the point estimates of the case fatality rate in the respective countries and the horizontal lines show the 95% confidence intervals. The open diamonds indicate pooled estimates by the fixed effect model and the black diamonds indicate pooled estimates by the random effect model.

Figure 2 shows the case number trajectories for newly diagnosed patients in the first 8 weeks of the epidemic in 78 countries. Assuming “independence” correlation structure, the GEE modeling showed significant time effect and time-by-group interaction. The coefficients of the GEE model can be translated so that the marginal mean rate ratio of the case increase in the non-BCG countries was 1.826 (95% CI 1.460 to 2.283, P=1×10^−6^) every week in initial 8 weeks. In BCG countries, the case increase rate was attenuated marginally by 25.4% (95% CI 3.0 to 42.7, P=0.029) as compared with those of the non-BCG countries.

**Figure 2:**
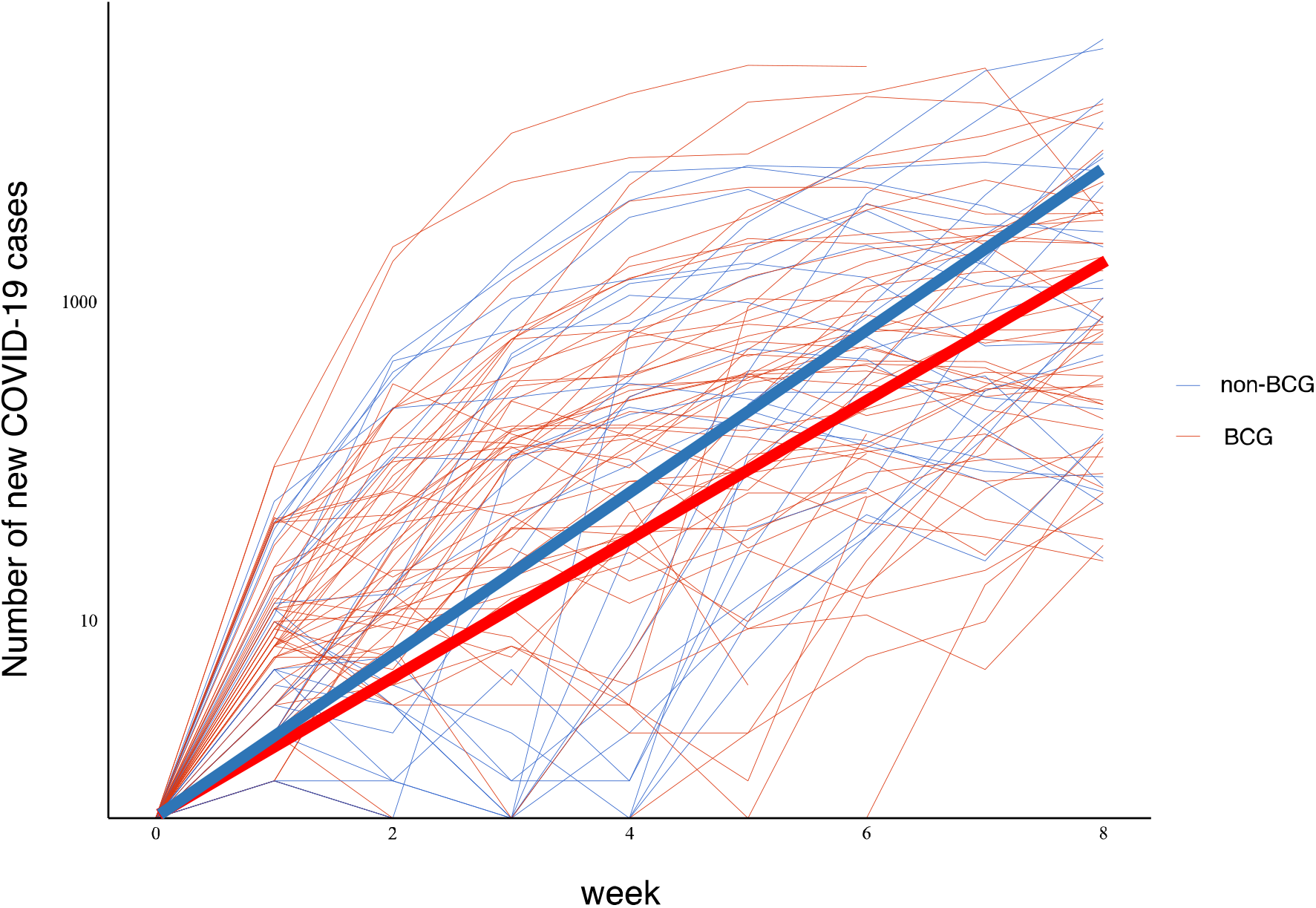
Time plot of newly diagnosed COVID-19 cases versus week for 78 countries. The bold lines indicate marginal mean trajectories of weekly new COVID-19 cases in a hypothetical country with 5,000,000 national population. Note that the y-axis is log-transformed.

## Discussion

My analysis indicates that countries with national mandatory BCG vaccination policies have lower reported COVID-19 mortality than those without such policies. Also, countries with current mandatory BCG policies show slower spread of COVID-19 infection than those without such policies. Although the protective mechanism of BCG vaccination against COVID-19 remains unknown, further laboratory and clinical research should be warranted.

## Data Availability

All data were extracted from publicly accessible databases. The sources of databases were shown in the references.

## References

1 Hegarty PK, Kamat A, Zafirakis H, Dinardo, A. BCG vaccination may be protective against Covid-19. Preprint. Posted online March, 2015. ResearchGate, doi:10.13140/RG.2.2.35948.10880.

2 Miller A, Reandelar MJ, Fasciglione K, Roumenova V, Li Y, Otazu GH. Correlation between universal BCG vaccination policy and reduced morbidity and mortality for COVID-19: an epidemiological study. Preprint. Posted online March 28, 2020. medRxiv doi:https://doi.org/10.1101/2020.03.24.20042937.

3 Akiyama Y, Ishida T. Relationship between COVID-19 death toll doubling time and national BCG vaccination policy. Preprint. Posted online April 21, 2020. medRxiv doi:10.1101/2020.04.06.20055251

4 John Hopkins University Coronavirus Resource Center. Coronavirus COVID-19 global cases. Accessed May 5, 2020. https://coronavirus.jhu.edu/map.html

5 The World Bank country classification by income 2020 fiscal year. Accessed April 30, 2020. https://datahelpdesk.worldbank.org/knowledgebase/articles/906519-world-bank-country-and-lending-groups

6 The World BCG Atlas, 2nd Edition. Accessed April 30, 2020. http://www.bcgatlas.org

